# Cost-utility analysis comparing ocrelizumab vs rituximab in the treatment of relapsing-remitting multiple sclerosis: The Colombian perspective

**DOI:** 10.1101/2022.08.23.22279145

**Authors:** Cristian Eduardo Navarro, John Edison Betancur, Alexandra Porras-Ramírez

## Abstract

**Introduction:** Since 2017, the ocrelizumab is available to treat patients with relapsing-remitting multiple sclerosis (RRMS), together with rituximab, they have a similar effectiveness but different costs. In this context, the added value provided by cost-effectiveness estimators for decision-making and drug prescription can be considered.

**Objective:** to determine the cost-utility of ocrelizumab versus rituximab in patients with RRMS, from the perspective of the Colombian health system.

**Methodology:** cost-utility study based on a Markov model, with a 50-year horizon and payer perspective. The currency was the US Dollar (USD) for the year 2019, with a threshold of $5,180 USD defined for Colombian health system. The model used annual cycles according to the health status determined by the disability scale. Direct costs were considered, and the incremental cost-effectiveness ratio (ICER) per 1 quality-adjusted life year (QALY) gained was used as the outcome measure. A discount rate of 5% was applied for costs and outcomes. Multiple one-way deterministic sensitivity analyzes and 10,000 modeling through Monte Carlo simulation were performed.

**Results:** for the treatment of patients with RRMS, ocrelizumab versus rituximab had an ICER of $73,652 USD for each QALY gained. After 50 years, 1 subject treated with ocrelizumab earns 4.8 QALYs more than 1 subject treated with rituximab, but at a higher cost of $521,759 USD vs $168,752 USD, respectively. Ocrelizumab becomes a cost-effective therapy when its price is discounted >86%, or there is a high willingness to pay.

**Conclusions:** Ocrelizumab was not a cost-effective drug compared with rituximab to treat patients with RRMS in Colombia.

## Introduction

Multiple sclerosis (MS) is a chronic, inflammatory and demyelinating disease of the central nervous system that leads to neuroaxonal degeneration ^1^, affecting females in a ratio of 3:1. The prevalence of MS in Colombia for the year 2016 was 5.52 cases per 100,000 inhabitants (2,662 total cases) ^2^, which is substantially lower than that of countries in other latitudes, such as England (112 × 100,000), Germany (85–118 × 100,000), Canada (55–248 × 100,000), USA (65–160 × 100,000) and Uruguay (30 × 100,000) ^3^. The male gender, age of >40 years, initial multisystemic involvement, frequent relapses, short interval between the first two relapses and a score of ≥4.0 on the Expanded Disability Status Scale (EDSS) are among the factors that are associated with a poor MS prognosis ^4,5^. Based on the above, there is a greater tendency of initiating an intensive therapy as early as possible if there is suspicion of poor prognosis ^6^. Almost all existing Disease-Modifying Therapies (DMTs) are available in Colombia, including ocrelizumab and rituximab; however, the latter is used off-label.

To the best of our knowledge, no head-to-head studies directly comparing the effectiveness of both anti-CD20 antibodies have been published to date. The phase 3 study by Christensen et al. ^7^ in Denmark should be completed in 2028 (ClinicalTrials.gov NCT04688788) and the phase 3 study by Torkildsen et al. ^8^ in Norway is scheduled for completion in 2025 (ClinicalTrials.gov NCT04578639). Two investigations not yet published in scientific journals (although available on the Internet) found that there is no difference between the two antibodies regarding the incidence of infusion-related reactions ^9,10^. A study analysing the Food and Drug Administration’s Adverse Event Reporting System (FAERS) database showed 7,948 and 623 reports for ocrelizumab and rituximab, respectively, with ocrelizumab having a stronger association with the incidence of infections compared with rituximab (21.93% vs 11.05%) ^11^.

MS is currently a high-cost disease for Colombia despite its low incidence. The cost varies depending on the DMT used, complications secondary to therapy, number of relapses, accumulated disability, and the need to conduct additional studies or hospitalize the patient. In 2014, the Colombian health insurance system spent $42,952,209 USD on patients with MS ^12^.

A cost-effectiveness study evaluating rituximab in relapsing–remitting MS (RRMS) has not been published to date ^13^; however, there have been several publications regarding ocrelizumab showing its cost-effectiveness compared with other DMTs ^14–17^. An article published in 2021 reported that the annual cost of rituximab in the US was $10,000 USD, while that of ocrelizumab was $65,000 USD ^18^, without considering the additional costs involved in drugs infusion, rehabilitation sessions, laboratory and diagnostic imaging tests, mobility aids and caregivers’ expenses. Hence, the money that a State must allocate to each patient with MS is incalculable ^19^.

The purpose of this investigation is to determine the cost-utility of ocrelizumab versus rituximab in patients with RRMS from the perspective of the Colombian health-care system.

## Methods

### Model structure (Figure 1)

A Markov model was designed to assess the cost-utility of ocrelizumab and rituximab for the treatment of patients with RRMS, with the following doses and infusion schedules:

Rituximab: first cycle, 1 g intravenous infusion on Day 1 and Day 15. Cycle repeated every 6 months, 1 g single-dose infusion.

**Figure 1:**
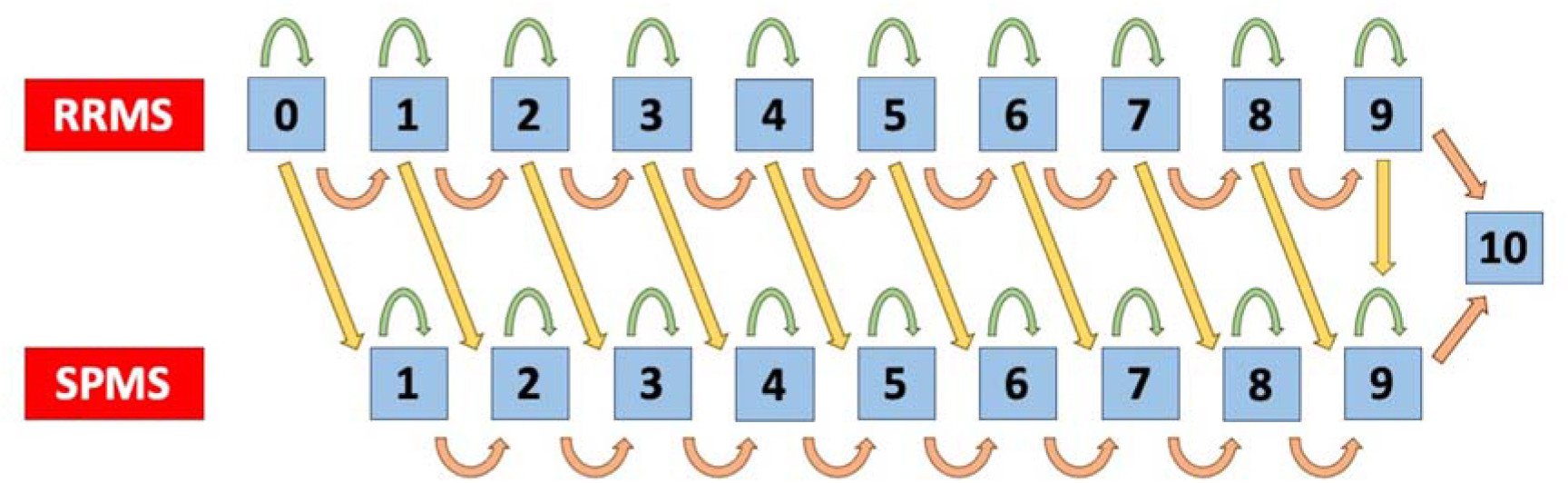
Structure of the model according to the Expanded Disability Status scale. EDSS: Expanded Disability Status Scale RRMS: Relapsing-Remitting Multiple Sclerosis SPMS: Secondary Progressive Multiple Sclerosis Green arrow: the patient remains in the same EDSS state of disability after 1 cycle Orange arrow: the patient moves to the next EDSS state of disability after 1 cycle Yellow arrow: the patient moves from RRMS to SPMS after 1 cycle

Ocrelizumab: first cycle, 300 mg intravenous infusion on Day 1 and Day 15. Cycle repeated every 6 months, 600 mg single-dose infusion.

This was based on the payer’s perspective in Colombia (General System of Social Security in Health) ^20,21^. The outcome of interest was the Incremental Cost-Effectiveness Ratio (ICER), which evaluated the cost gained per 1 Quality-Adjusted Life Year (QALY). A 5% discount rate was applied for the costs and outcomes to do the model comparable with other studies published and considering the recommendations of *Instituto de Evaluación Tecnológica en Salud* (IETS) for Colombia.

### Data processing and assumptions for the development of the model Target population

A theoretical cohort of 1,000 patients with RRMS and an EDSS score 0 were included in the model, and the therapy was started with either ocrelizumab or rituximab. The horizon considered was 50 years, according to the life expectancy at birth in Colombia. The model considered 21 health statuses (EDSS 0–9 in RRMS, EDSS 0–9 in secondary progressive MS [SPMS] and EDSS 10 for death), with cycles occurring annually. The health status was defined with 1-point increments on the EDSS scale. The transition probabilities between each status were extracted from the natural history studies of the disease conducted by Palace et al. ^22^ and Huygens et al. ^23^ (Appendix A Table 1 in Supplemental materials). Data regarding the effectiveness of each therapy (Incidence Rate Ratio [IRR]) and mortality (Hazard Ratio [HR]) were extracted for each EDSS status from this latest publication.

**Table 1.**
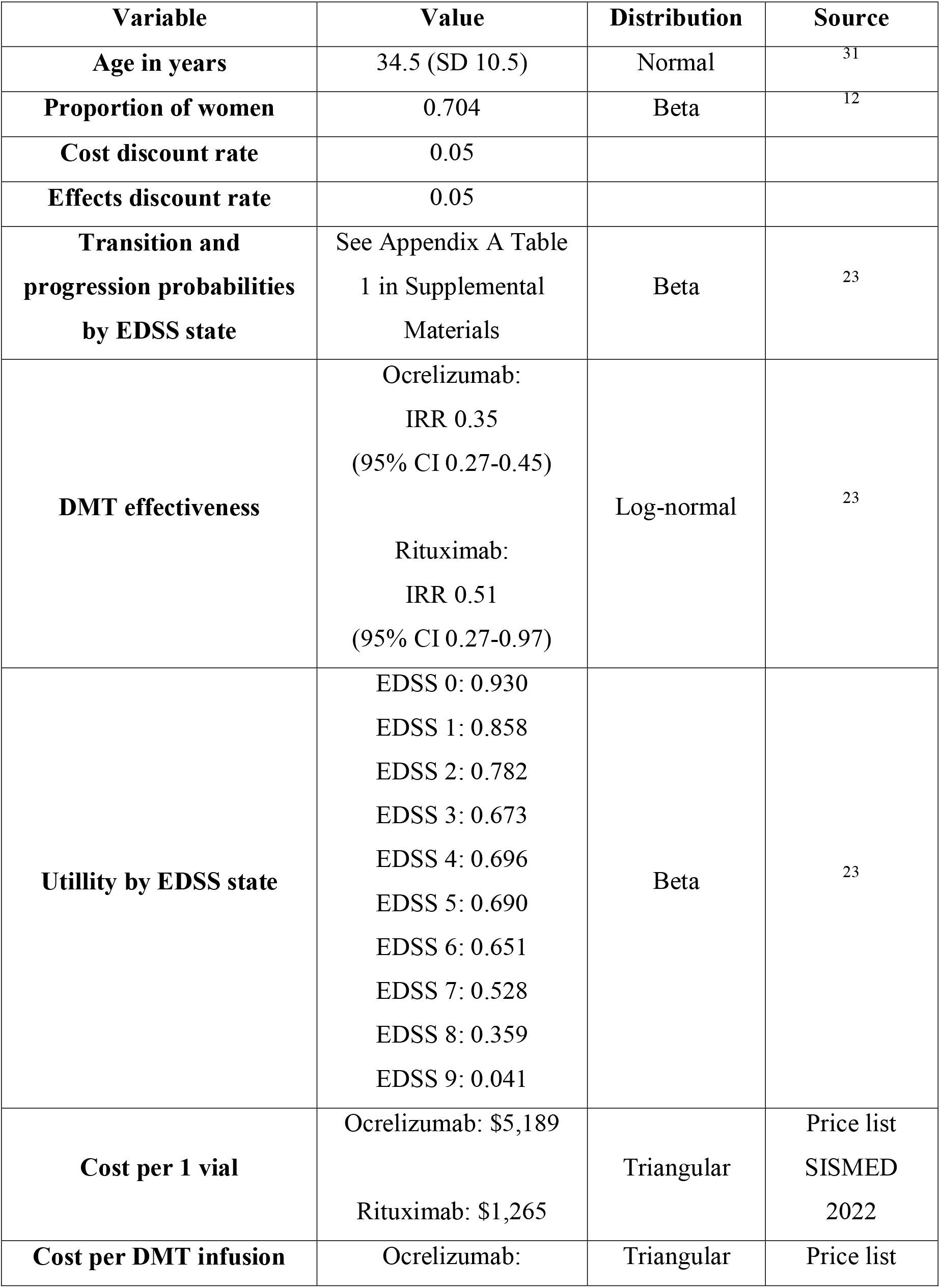
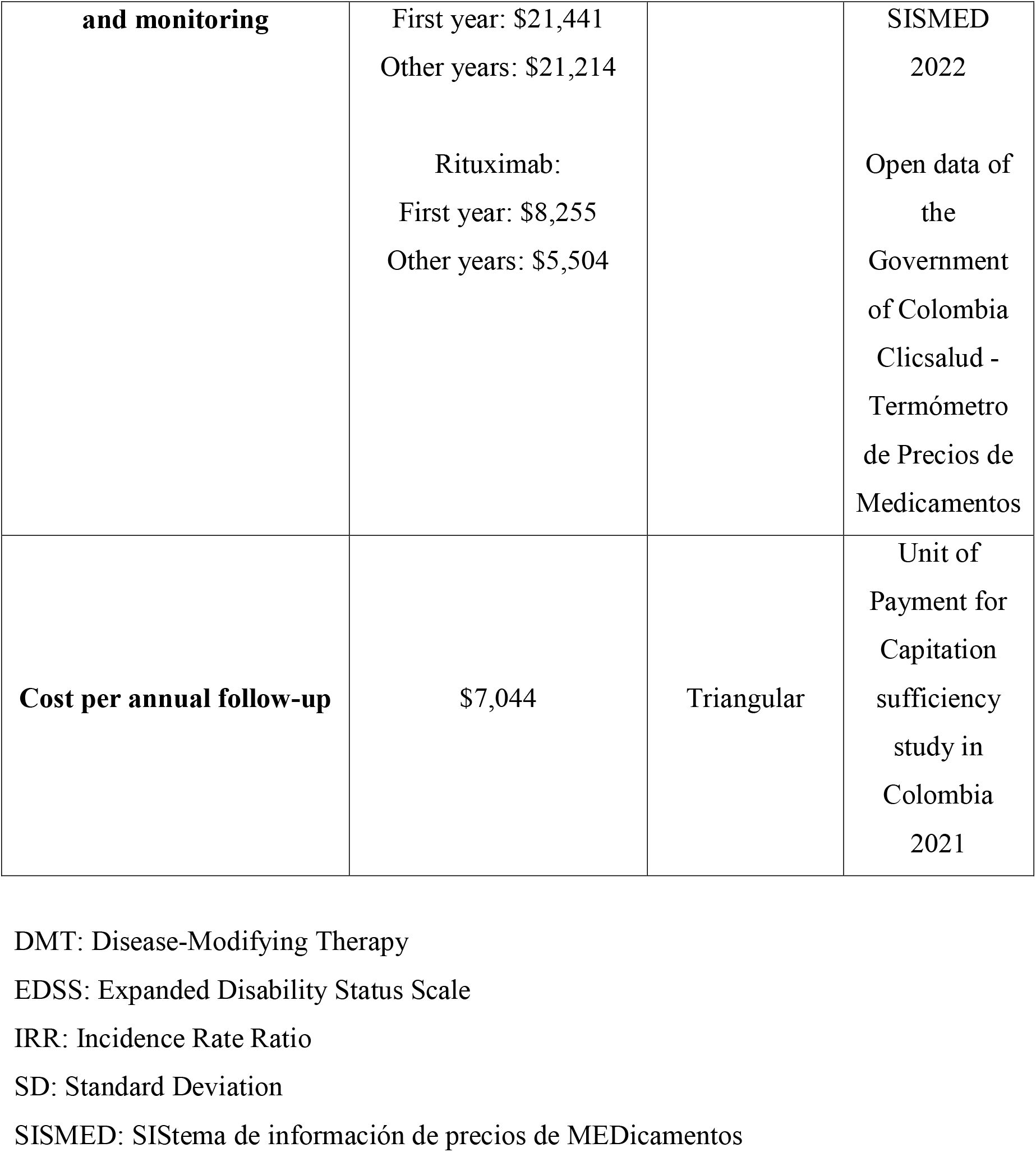
Summary of model input parameters

The pharmacological interventions did not change the transition probabilities from RRMS to SPMS. Patients entered the model without disability (EDSS 0) and moved between the statuses in each cycle according to the probabilities. They could be in the RRMS stage or move on to the SPMS stage, ending up in the absorbing status at the end (EDSS 10 or death by MS). Treatment was discontinued when a subject reached a RRMS EDSS status of ≥7 or transitioned to any SPMS status (no anti-CD20 antibody is authorized for that stage). The model did not consider the use of another DMT a posteriori to avoid a confounding factor.

### Effects and utilities

The rituximab and ocrelizumab IRRs were extracted from a recent network meta-analysis ^23^. The capacity of the DMTs to arrest disability progression was modelled by applying the IRR to the EDSS transition probabilities. The IRR was independent of the EDSS score, which indicated that the effect of therapy was not lost over time. The model used the most up-to-date information known regarding the disease’s natural history ^22,23^.

The utilities according to the EDSS scale were extracted from the study conducted by Huygens et al. ^23^, which were based on the EQ-5D. No additional decrease in utility was applied if the subject was at the SPMS stage. The utilities determined in the study by Orme et al. ^24^ were not chosen since the measurement of disability was conducted using the Adapted Patient Determined Disease Steps scale (APDDS), which differs from the EDSS in its scores (6.5 to10) ^25^.

### Costs

The spending resources of monoclonal antibody administration and monitoring disability were identified with the advice of two experts in MS (a head nurse and fellow neurologist in MS who are the coordinators of the *Centro de Esclerosis Múltiple del Hospital Universitario Nacional* [CEMHUN] in Bogotá). The costs were estimated using the fees of the 2022 SISMED *(SIStema de información de precios de MEDicamentos)*, the fees of the open data page of the Government of Colombia ^26^, and the 2022 Unit of Payment for Capitation sufficiency study. A base-case was modelled with these results in which the costs of one and 50 years of treatment per patient with each drug were determined.

The threshold used was $5,180 USD, which was defined by IETS for the Colombian health system ^27^. The market exchanged rate for the year 2019 reported by Banco de la República ^28^ ($1 USD = $3,281 COP) was employed to perform the conversion and estimated adjustment of the costs to the value of the dollar for the year 2019.

#### Direct costs considered

Cost per one vial of DMT: expense in the acquisition of one vial of the drug.

Cost per administration of treatment: expense in the administration of DMT, including the medical supplies and pre-medication required, and costs related to the monitoring of adverse reaction incidences and their management. The infusion room, doctor and nursing expenses were included in this value.

Cost of annual follow-up: expenses for consultation with specialists (neurologist, psychiatrist, physiatrist, psychologist), cognitive evaluation, serum creatinine test, annual contrasted brain and spinal cord magnetic resonance imaging, rehabilitation (physical, occupational, speech therapy) and vitamin D supplement for 1 year. It was assumed that no additional follow-up studies with these anti-CD20 monoclonal antibodies were required. Total cost of the disease per 1 year: cost of DMT, its administration and follow-up for one patient per 1 year.

Total cost of the disease per patient: cost of DMT, its administration and follow-up for one patient over a 50-year horizon. The infusion scheme was considered and the patients were assumed to be 100% adherent.

The summary of the input parameters of the model is shown in Table 1.

### Sensitivity analysis

The sensitivity analysis included a univariate deterministic analysis where the individual effect of different model parameters was assessed. The evaluated parameters were: discount rate (0%, 3.5%, 7% and 12%), DMT costs (+/−30%), DMT administration and monitoring costs (+/−30%), annual follow-up costs (+/−30%), IRR of each DMT (according to 95% confidence interval [CI]), utilities (+/−30%), time horizon (20, 30 and 100 years) and non-discontinuation of DMT during all RRMS stage disease. The other type of sensitivity analysis performed was probabilistic, performing 10,000 models through Monte Carlo simulation and modifying the costs (+/−30%) and QALYs (+/−30%). This analysis runs through the probability distributions of each variable.

Microsoft Excel - Office 365^®^ software was used to save the information and run the model.

### Bias type and control

Bias mitigation was performed based on the recommendation of Evers et al. ^29^ and is included in Appendix B in Supplemental materials.

## Results

### Base-case scenario

According to the model proposed for the Colombian context, ocrelizumab compared with rituximab in the treatment of patients with RRMS had an ICER of $73,652 USD for each QALY gained ($5,180 USD threshold).

One vial of ocrelizumab costs four times the price of one vial of rituximab ($5,189 USD vs. $1,265 USD). The cost of ocrelizumab administration during the first year was 2.6 times the cost of rituximab administration ($21,441 USD vs $8,255 USD). This cost remains very similar in the future cycles ($21,214 USD); however, the cost of rituximab administration in the following years is lowered by 33% compared with the initial cycle ($5,504 USD). If a patient continues in RRMS state with a EDSS <6, ocrelizumab therapy over a 50-year horizon costs three times that of rituximab therapy (after applying a 5% discount rate). Regarding the QALYs gained, after 50 years, one subject treated with ocrelizumab gained 4.8 more QALYs than a subject treated with rituximab (Table 2).

**Table 2.** Summary of base-case scenario results

| **Variable** | **Ocrelizumab** | **Rituximab** | **Incremental** |
| --- | --- | --- | --- |
| Cost per 1 patient per 50 years | \$521,759 | \$168,752 | \$353,007 |
| QALYs per 1 patient per 50 years | 16.44 | 11.65 | 4.79 |
| ICER | NA | NA | \$73,652 |

ICER: Incremental Cost-Effectiveness Ratio
QALY: Quality-Adjusted Life-Years
NA: Not Applicable
Threshold = \$5,180 USD. Market exchanged rate 2019: \$1 USD = \$3,281 COP

There were no differences in the costs of the administration and monitoring of each DMT since they are similar therapies in terms of pre-medication and their six-monthly application.

### Univariate deterministic sensitivity analysis

Ocrelizumab continued to be non-cost–effective compared with rituximab in each of the scenarios considered. Appendix C Table 2 in Supplemental materials shows the input parameters of the model. The variable with the most significant impact on the ICER was the change in the IRR of ocrelizumab (from $54,868 USD to $169,387 USD per QALY), followed by an intervention horizon of 20 years ($106,254 USD per QALY), and in third place was the cost of one vial of ocrelizumab (from $49,882 USD to $97,423 USD per QALY). The variable with the least impact on the change in the ICER value was the administration of DMT during all the stages of RRMS ($73,653 USD per QALY). The change of the IRR of rituximab was found to make the therapy dominant over ocrelizumab (-$58,252 USD per QALY) (Figure 2 and Appendix D Table 3 in Supplemental materials).

**Figure 2:**
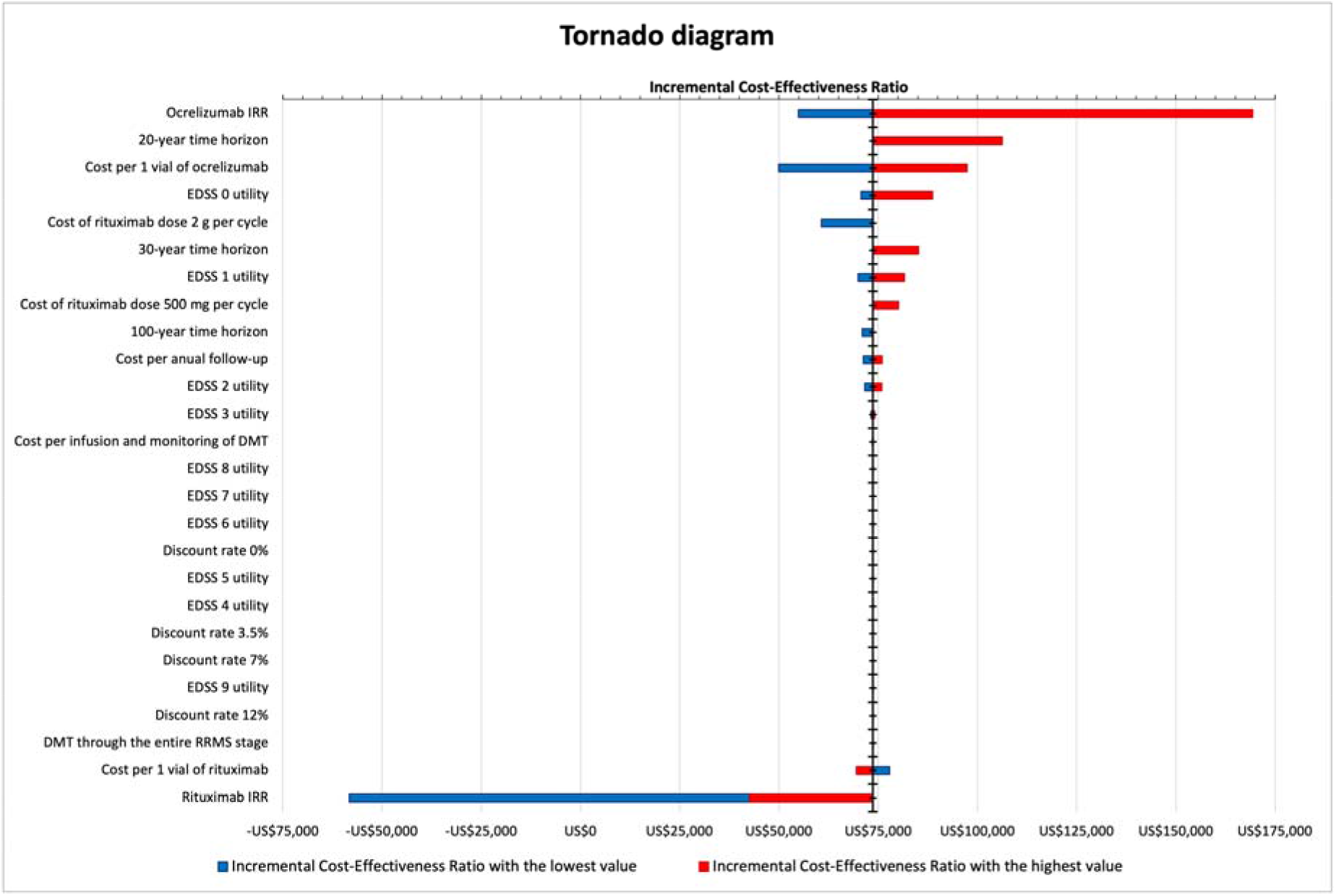
Tornado diagram of the one-way deterministic sensitivity analysis DMT: Disease-Modifying Therapy EDSS: Expanded Disability Status Scale IRR: Incidence Rate Ratio RRMS: Relapsing-Remitting Multiple Sclerosis

Initially, we had planned to modify the cost of DMT +/−30%; however, considering that ocrelizumab was well above the threshold, we decided to conduct a new one-way sensitivity analysis by modifying the cost of one vial of this drug from 100% to 10% of its current value. The result is presented in the graph of ICER versus cost (Figure 3). Ocrelizumab becomes a cost-effective therapy when its price is below 14% of its current value (<$727 USD per 1 vial), that is, when a >86% discount is applied (Appendix E Table 4 in Supplemental materials).

**Figure 3.**
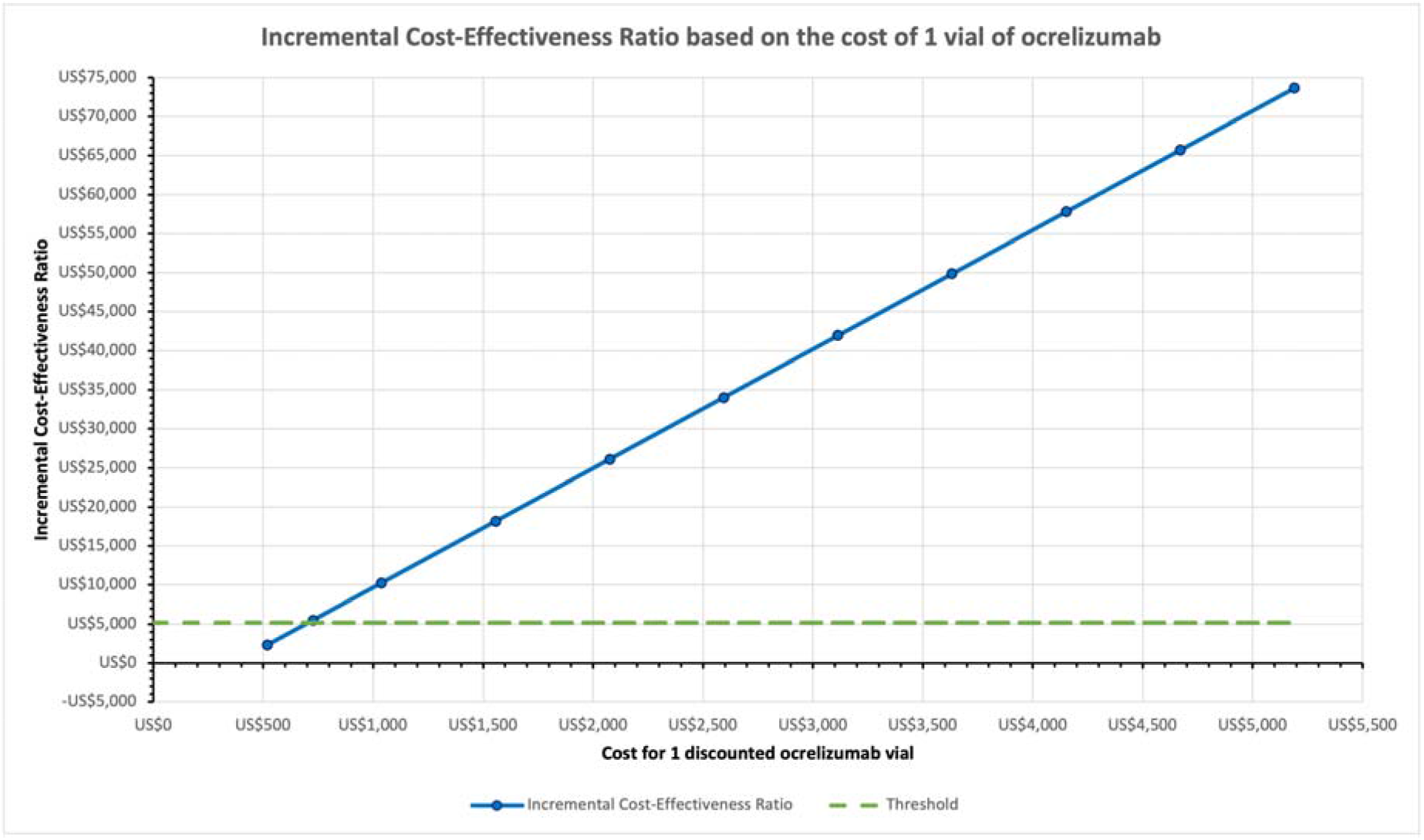
Incremental Cost-Effectiveness Ratio based on the cost of 1 vial of ocrelizumab

### Probabilistic sensitivity analysis

Rituximab was dominant over ocrelizumab in 29.8% of the simulations (Appendix F Figure 1 in Supplemental materials). Ocrelizumab did not have an ICER <$5,180 USD in any of the simulated scenarios (Figure 4).

**Figure 4.**
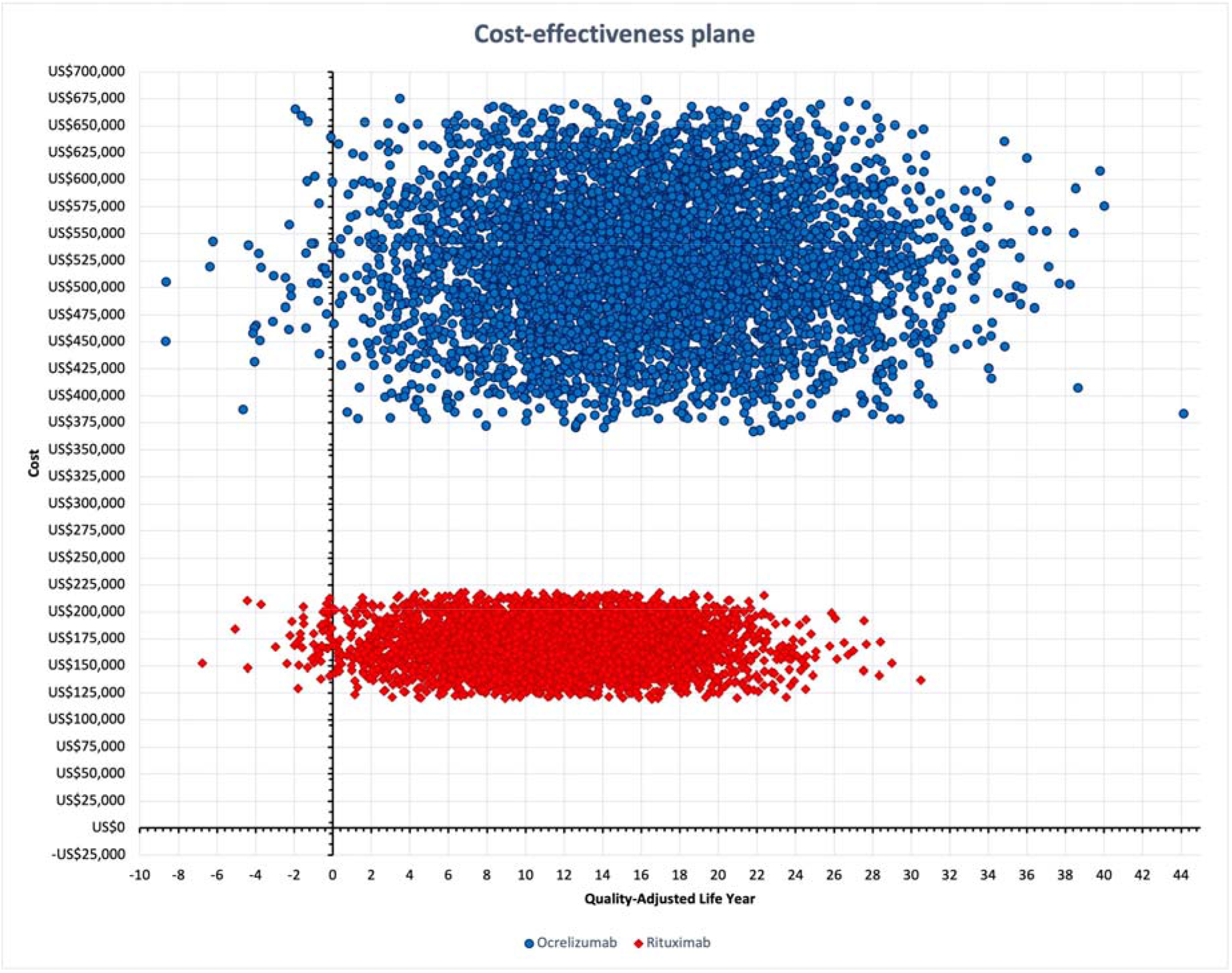
Scatter plot of the probabilistic sensitivity analysis of both disease-modifying therapies.

For ocrelizumab to have a probability of being cost-effective by at least 50%, the willingness to pay must be $80,000 USD (Appendix G Figure 2 in Supplemental materials).

## Discussion

The model in this study, generated from the perspective of the payer in Colombia, showed that ocrelizumab is not a cost-effective therapy for the treatment of patients with RRMS with a horizon of 50 years. In other words, ocrelizumab increases the total costs without generating a significant increase in QALYs compared with the benefit generated by rituximab.

The results mentioned above are robust and were obtained in the different scenarios proposed for both the types of sensitivity analyses. The primary reason for this is based on the cost difference between both the DMTs. A variability range of +/−30% in the costs of one vial of each drug was proposed and the limits of the interval did not overlap despite this, as shown in Appendix C Table 2 in Supplemental materials. The ICER of ocrelizumab decreases when used a 50-year horizon, since the ICER increases from $73,652 USD to $106,254 USD when used a 20-year horizon (difference of +$32,602 USD). However, when a 100-year horizon is considered, there is no greater discount over time (ICER $70,934 USD; difference of -$2,719 USD).

Moreover, the cost of ocrelizumab is so high that the discount rates do not impact the ICER, as shown in the tornado diagram (Figure 2). Ocrelizumab becomes cost-effective for the Colombian context only when its price drops to less than 14% of its current value (<$727 USD per vial) or the willingness to pay increases markedly, which would also increase health spending remarkably.

Another explanation for the lack of cost-effectiveness of ocrelizumab is the lack of greater benefit, since the 95% CI of the IRR of each DMT overlaps (ocrelizumab IRR 0.35 [95% CI 0.27–0.45] vs rituximab IRR 0.51 [95% CI 0.27–0.97]). This indicates that ocrelizumab is not more effective than rituximab in RRMS treatment. In the 10,000 Monte Carlo simulations, rituximab turned out to be dominant over ocrelizumab in 29.8% of the scenarios, as it generated greater benefit in QALYs, which is in line with the abovementioned data.

The dose from the second cycle of rituximab drops from 2 g to 1 g, which contributes to less expenditure on its administration. Considering that some studies have evaluated the effectiveness of rituximab with the dose of 2 g vs 1 g vs 500 mg, deterministic sensitivity analysis did not show that using a higher dose of rituximab (and therefore increasing the cost) generated a significant change in the ICER in favour of ocrelizumab ($60,686 USD), which is still far from the threshold for Colombia.

This investigation is the first known cost-utility study that compares two anti-CD20 antibodies used in the treatment of patients with RRMS. The result mentioned in the pharmacoeconomic report published in 2017 by the Canadian Agency for Drugs and Technologies in Health is consistent with that obtained in this study ^30^. The analysis differs slightly in the horizon, since the drug was evaluated for up to 63 years, but the comparison was made with the other DMTs available in Canada. The major difference is the $50,000 CAD threshold, but despite this, the probability of being cost-effective was only 2%.

Yang H et al. ^14^ documented in 2017 that ocrelizumab is cost-effective in almost all the scenarios proposed, but when it was compared with a less effective therapy such as interferon beta-1a, using a threshold of $100,000 USD. Finally, the investigation by Chirokov et al. ^17^ considered the loss of effectiveness of DMT as the cycles pass through the horizon, a detail that was not considered in this study and would further contribute to increasing the ICER against ocrelizumab. This last study used a 20-year horizon, documenting the costs of $908,365 USD and 8,49 QALYs for ocrelizumab, which differ from the $395,178 USD and 12,54 QALYs found in the scenario with a 20-year horizon by deterministic sensitivity analysis in this investigation. This is due to the price of the drug and the transition probabilities and utilities used, since the IRR of 0.35 was the same in both the studies. Considering this, there is little comparability of the analyses due to their heterogeneous methodology, which prevents the extrapolation of the results to a local context ^13^. Therefore, it becomes imperative to present these cost-utility analyses from the Colombian perspective.

The results obtained provide information to determine which anti-CD20 monoclonal antibody available in Colombia for RRMS treatment is not cost-effective. Unfortunately, ocrelizumab is the only anti-CD20 antibody that has been approved in Colombia by a regulatory entity to be used as a treatment against RRMS, generating excessive (and perhaps unnecessary) expense for the health insurance system, especially since rituximab is available and generates similar benefits at a lower cost. We expect that the data obtained in this study will favour a better and more rational use of public health resources and allow a better distribution of expenditure and savings. Further, this will promote more strategies aimed at rehabilitating patients with MS, considering the economic impact that can be achieved and alleviating the budgetary burden of a medical problem that is chronic and disabling.

A prescription guideline taking into consideration the economic factor can be generated, but the regulatory entities are required to facilitate the use of rituximab to treat this disease with the available clinical evidence. The above is necessary since the development of new molecules that involve novel technology will otherwise continue without providing greater effectiveness, but rapidly exhausting public money.

The limitations of this study include the fact that the IRR values of each DMT were not obtained from the phase 3 studies that compare them ‘head-to-head,’ instead they were obtained from the estimates made in the most recent network meta-analysis published ^23^. In turn, the natural history data used to obtain the transition probabilities originate from the best adjusted model published to date ^22,23^. Thus, the results can vary markedly according to the origin of these data. We hope that this model will be replicated when the results of the still-ongoing phase 3 studies that compare ocrelizumab with rituximab in RRMS become available ^7,8^. Another limitation is that although the model tries to adjust as well as possible, it does not reflect the reality of each patient individually, since a patient can move between each cycle more slowly or quickly than another or can even improve their disability and reduce their EDSS score. Furthermore, the model did not consider the use of subsequent therapies for the SPMS stage, such as the use of siponimod. Finally, information regarding how much money is saved per relapse was not presented ^14,17^, since there was no difference between their effectiveness.

## Conclusions

Ocrelizumab is not a cost-effective therapy when compared with rituximab if there is a willingness to pay of $5,180 USD per QALY gained. For ocrelizumab to be cost-effective, one vial must be worth less than $727 USD. Ocrelizumab has a 50% chance of being cost-effective whether the health-care system pays at least $80,000 USD per QALY earned. Rituximab was dominant over ocrelizumab in 29.8% of the simulations performed in this study.

Therefore, rituximab appears to be an equally effective and less expensive alternative for treating patients with RRMS in Colombia. We propose that decision-makers consider rituximab as a cost-effective therapy for RRMS. Situations such as the one presented in this article occur in different high-cost or orphan diseases; hence, it is imperative to conduct pharmacoeconomic studies to determine the actual usefulness of expensive therapies or interventions in specific population groups.

## Supporting information

CHEERS checklist

## Data Availability

All data produced in the present work are contained in the manuscript

## Declarations

### Funding

This research did not receive any specific grant from funding agencies in the public, commercial, or not-for-profit sectors.

### Conflicts of Interest

Cristian Eduardo Navarro, John Edison Betancur, Alexandra Porras-Ramírez declare that they have no conflict of interest.

### Ethical approval

Ethics approval is not required for health economic assessment at our institution.

### Consent to participate and for publication

Not required

### Availability of data and material

The datasets generated during and/or analyzed during the current study are not publicly available as they contain proprietary data but are available from the corresponding author on reasonable request.

### Author contributions

Conceptualization: Cristian Eduardo Navarro; Methodology: Cristian Eduardo Navarro, John Edison Betancur, Alexandra Porras-Ramírez; Formal analysis and investigation: Cristian Eduardo Navarro; Writing - original draft preparation: Cristian Eduardo Navarro; Writing - review and editing: Cristian Eduardo Navarro, John Edison Betancur, Alexandra Porras-Ramírez; Resources: Cristian Eduardo Navarro; Supervision: John Edison Betancur.

## Acknowledgements

To Lorena López and Simón Cárdenas, who coordinate the Centro de Esclerosis Múltiple del Hospital Universitario Nacional (CEMHUN) in Colombia, and who helped us with the costing of the interventions.

## Notes

### Competing Interest Statement

The authors have declared no competing interest.

### Funding Statement

This study did not receive any funding

