## Supplemental materials for "Cost-utility analysis comparing ocrelizumab vs rituximab in the treatment of relapsing-remitting multiple sclerosis: The Colombian perspective"

1. Unit of Clinical Neurology, School of Medicine, Universidad Nacional de Colombia. Bogotá, Colombia.
2. Master's program in Epidemiology. School of Medicine, Universidad El Bosque. Bogotá, Colombia.
3. Grupo de Investigación en Neurología de la Universidad Nacional de Colombia – NeuroUnal. Bogotá, Colombia
4. School of Medicine, Universidad de Antioquia. Medellín, Colombia.

*** Correspondence and reprints:**

Cristian Eduardo Navarro MD MSc

Unit of Clinical Neurology, School of Medicine, Universidad Nacional de Colombia.

Address: Ciudad Universitaria carrera 30 # 45-03. Bogotá, Colombia.

Postal code: 111321

Cellphone: (+57) 3004730287

https://orcid.org/0000-0003-0532-6301

**Appendix A.** Table 1. Transition probabilities according to the natural history of the disease (continue below).

| | | RRMS | | | | | | | | | |
|---|---|---|---|---|---|---|---|---|---|---|---|
| | | **EDSS0** | **EDSS1** | **EDSS2** | **EDSS3** | **EDSS4** | **EDSS5** | **EDSS6** | **EDSS7** | **EDSS8** | **EDSS9** |
| **RRMS** | **EDSS0** | 0.695 | 0.203 | 0.073 | 0.022 | 0.004 | 0.001 | 0.002 | 0 | 0 | 0 |
| | **EDSS1** | 0.058 | 0.693 | 0.157 | 0.061 | 0.016 | 0.005 | 0.006 | 0 | 0 | 0 |
| | **EDSS2** | 0.016 | 0.121 | 0.583 | 0.161 | 0.045 | 0.018 | 0.022 | 0.002 | 0.001 | 0 |
| | **EDSS3** | 0.006 | 0.050 | 0.120 | 0.444 | 0.074 | 0.058 | 0.116 | 0.010 | 0.004 | 0 |
| | **EDSS4** | 0.002 | 0.022 | 0.067 | 0.115 | 0.390 | 0.083 | 0.168 | 0.026 | 0.007 | 0.001 |
| | **EDSS5** | 0.001 | 0.005 | 0.029 | 0.059 | 0.087 | 0.295 | 0.166 | 0.039 | 0.019 | 0.001 |
| | **EDSS6** | 0 | 0.001 | 0.004 | 0.025 | 0.031 | 0.041 | 0.534 | 0.079 | 0.044 | 0.004 |
| | **EDSS7** | 0 | 0 | 0.001 | 0.002 | 0.007 | 0.004 | 0.117 | 0.486 | 0.113 | 0.016 |
| | **EDSS8** | 0 | 0 | 0 | 0 | 0.001 | 0.001 | 0.019 | 0.056 | 0.754 | 0.017 |
| | **EDSS9** | 0 | 0 | 0 | 0 | 0 | 0 | 0.002 | 0.006 | 0.174 | 0 |
| **SPMS** | **EDSS1** | 0 | 0 | 0 | 0 | 0 | 0 | 0 | 0 | 0 | 0 |
| | **EDSS2** | 0 | 0 | 0 | 0 | 0 | 0 | 0 | 0 | 0 | 0 |
| | **EDSS3** | 0 | 0 | 0 | 0 | 0 | 0 | 0 | 0 | 0 | 0 |
| | **EDSS4** | 0 | 0 | 0 | 0 | 0 | 0 | 0 | 0 | 0 | 0 |
| | **EDSS5** | 0 | 0 | 0 | 0 | 0 | 0 | 0 | 0 | 0 | 0 |
| | **EDSS6** | 0 | 0 | 0 | 0 | 0 | 0 | 0 | 0 | 0 | 0 |
| | **EDSS7** | 0 | 0 | 0 | 0 | 0 | 0 | 0 | 0 | 0 | 0 |
| | **EDSS8** | 0 | 0 | 0 | 0 | 0 | 0 | 0 | 0 | 0 | 0 |
| | **EDSS9** | 0 | 0 | 0 | 0 | 0 | 0 | 0 | 0 | 0 | 0 |

EDSS: Expanded Disability Status Scale

RRMS: Relapsing-Remitting Multiple Sclerosis

SPMS: Secondary Progressive Multiple Sclerosis

**Appendix A.** Table 1. Transition probabilities according to the natural history of the disease (continuation).

| | | SPMS | | | | | | | | |
|---|---|---|---|---|---|---|---|---|---|---|
| | | **EDSS1** | **EDSS2** | **EDSS3** | **EDSS4** | **EDSS5** | **EDSS6** | **EDSS7** | **EDSS8** | **EDSS9** |
| **RRMS** | **EDSS0** | 0 | 0 | 0 | 0 | 0 | 0 | 0 | 0 | 0 |
| | **EDSS1** | 0 | 0.003 | 0 | 0 | 0 | 0 | 0 | 0 | 0 |
| | **EDSS2** | 0 | 0 | 0.032 | 0 | 0 | 0 | 0 | 0 | 0 |
| | **EDSS3** | 0 | 0 | 0 | 0.117 | 0 | 0 | 0 | 0 | 0 |
| | **EDSS4** | 0 | 0 | 0 | 0 | 0.120 | 0 | 0 | 0 | 0 |
| | **EDSS5** | 0 | 0 | 0 | 0 | 0 | 0.299 | 0 | 0 | 0 |
| | **EDSS6** | 0 | 0 | 0 | 0 | 0 | 0 | 0.237 | 0 | 0 |
| | **EDSS7** | 0 | 0 | 0 | 0 | 0 | 0 | 0 | 0.254 | 0 |
| | **EDSS8** | 0 | 0 | 0 | 0 | 0 | 0 | 0 | 0 | 0.153 |
| | **EDSS9** | 0 | 0 | 0 | 0 | 0 | 0 | 0 | 0 | 0.818 |
| **SPMS** | **EDSS1** | 0.769 | 0.154 | 0.077 | 0 | 0 | 0 | 0 | 0 | 0 |
| | **EDSS2** | 0 | 0.636 | 0.271 | 0.062 | 0.023 | 0.008 | 0 | 0 | 0 |
| | **EDSS3** | 0 | 0 | 0.629 | 0.253 | 0.077 | 0.033 | 0.003 | 0.005 | 0 |
| | **EDSS4** | 0 | 0 | 0 | 0.485 | 0.350 | 0.139 | 0.007 | 0.018 | 0 |
| | **EDSS5** | 0 | 0 | 0 | 0 | 0.633 | 0.317 | 0.022 | 0.026 | 0.002 |
| | **EDSS6** | 0 | 0 | 0 | 0 | 0 | 0.763 | 0.190 | 0.045 | 0.002 |
| | **EDSS7** | 0 | 0 | 0 | 0 | 0 | 0 | 0.805 | 0.189 | 0.006 |
| | **EDSS8** | 0 | 0 | 0 | 0 | 0 | 0 | 0 | 0.926 | 0.074 |
| | **EDSS9** | 0 | 0 | 0 | 0 | 0 | 0 | 0 | 0 | 1.000 |

EDSS: Expanded Disability Status Scale

RRMS: Relapsing-Remitting Multiple Sclerosis

SPMS: Secondary Progressive Multiple Sclerosis

**Appendix B.** Types and bias control

Bias control was performed considering the Evers et al's recommendations [1]:

- Pre-trial bias:
  - Narrow perspective bias: it could not be avoided; the IETS recommendation for Colombia is to carry out studies with a payer perspective that is narrower and omits other types of costs.
  - Inefficient comparator bias: two similarly effective therapies were compared.
  - Cost measurement omission bias: cost that can generate a negative effect on the cost-effectiveness of therapies were included.
  - Intermittent data collection bias: does not apply to the Markov model.
- Bias during trial:
  - Invalid valuation bias: the correct monetary value was given to each measurement.
  - Ordinal ICER bias: does not apply because ordinal scales are not used in the ICER calculation.
  - Double-counting bias: it was avoided by not considering the same cost more than once.
  - Inappropriate discounting bias: IETS recommendations for Colombia were considered.
  - Limited sensitivity analysis bias: IETS recommendations for Colombia were taken into account.
- Bias after trial:
  - Sponsor bias: the study had no sponsor.
  - Reporting and dissemination bias: regardless of the results, the study will be published.

**Appendix C.** Table 2. Input parameters of the univariate deterministic sensitivity analysis.

| Variable | Lowest value | Base-case value | Highest value |
|---|---|---|---|
| Cost per 1 vial of ocrelizumab | $3,633 | $5,189 | $6,746 |
| Cost per 1 vial of rituximab | $886 | $1,265 | $1,645 |
| Cost per infusion and monitoring of DMT | $159 | $228 | $296 |
| Cost per annual follow-up | $4,931 | $7,044 | $9,157 |
| EDSS 0 utility | 0.651 | 0.93 | 1 |
| EDSS 1 utility | 0.6006 | 0.858 | 1 |
| EDSS 2 utility | 0.5474 | 0.782 | 1 |
| EDSS 3 utility | 0.4711 | 0.673 | 0.8749 |
| EDSS 4 utility | 0.4872 | 0.696 | 0.9048 |
| EDSS 5 utility | 0.483 | 0.69 | 0.897 |
| EDSS 6 utility | 0.4557 | 0.651 | 0.8463 |
| EDSS 7 utility | 0.3696 | 0.528 | 0.6864 |
| EDSS 8 utility | 0.2513 | 0.359 | 0.4667 |
| EDSS 9 utility | 0.0287 | 0.041 | 0.0533 |

DMT: Disease-Modifying Therapy

EDSS: Expanded Disability Status Scale

Threshold = $5,180 USD. Market exchanged rate 2019: $1 USD = $3,281 COP

**Appendix D.** Table 3. Results of the univariate deterministic sensitivity analysis

| Variable | ICER with the lowest value | ICER base-case | ICER with the highest value |
|---|---|---|---|
| Ocrelizumab IRR | $54,868 | $73,652 | $169,387 |
| 20-year time horizon | | $73,652 | $106,254 |
| Cost per 1 vial of ocrelizumab | $49,882 | $73,652 | $97,423 |
| EDSS 0 utility | $70,649 | $73,652 | $88,676 |
| Cost of rituximab dose 2 g per cycle | $60,686 | $73,652 | |
| 30-year time horizon | | $73,652 | $85,139 |
| EDSS 1 utility | $69,895 | $73,652 | $81,605 |
| Cost of rituximab dose 500 mg per cycle | | $73,652 | $80,135 |
| 100-year time horizon | $70,934 | $73,652 | |
| Cost per anual follow-up | $71,288 | $73,652 | $76,017 |
| EDSS 2 utility | $71,647 | $73,652 | $75,940 |
| EDSS 3 utility | $73,189 | $73,652 | $74,122 |
| Cost per infusion and monitoring of DMT | $73,462 | $73,652 | $73,776 |
| EDSS 8 utility | $73,539 | $73,652 | $73,766 |
| EDSS 7 utility | $73,590 | $73,652 | $73,715 |
| EDSS 6 utility | $73,593 | $73,652 | $73,712 |
| Discount rate 0% | $73,596 | $73,652 | |
| EDSS 5 utility | $73,623 | $73,652 | $73,682 |
| EDSS 4 utility | $73,629 | $73,652 | $73,676 |
| Discount rate 3.5% | $73,646 | $73,652 | |
| Discount rate 7% | | $73,652 | $73,655 |
| EDSS 9 utility | $73,650 | $73,652 | $73,655 |
| Discount rate 12% | | $73,652 | $73,653 |

| | | | |
|---|---|---|---|
| DMT through the entire RRMS stage | | $73,652 | $73,653 |
| Cost per 1 vial of rituximab | $77,859 | $73,652 | $69,446 |
| Rituximab IRR | -$58,252 | $73,652 | $42,537 |

DMT: Disease-Modifying Therapy

EDSS: Expanded Disability Status Scale

ICER: Incremental Cost-Effectiveness Ratio

IRR: Incidence Rate Ratio

RRMS: Relapsing-Remitting Multiple Sclerosis

Threshold = $5,180 USD. Market exchanged rate 2019: $1 USD = $3,281 COP

**Appendix E.** Table 4. Discount value per 1 vial of ocrelizumab and its associated ICER.

| Discount | Cost for 1 discounted ocrelizumab vial | ICER |
|---|---|---|
| 0% | $5,189 | $73,652 |
| 10% | $4,671 | $65,729 |
| 20% | $4,152 | $57,805 |
| 30% | $3,633 | $49,882 |
| 40% | $3,114 | $41,958 |
| 50% | $2,595 | $34,034 |
| 60% | $2,076 | $26,111 |
| 70% | $1,557 | $18,187 |
| 80% | $1,038 | $10,264 |
| 86% | $727 | $5,510 |
| 90% | $519 | $2,340 |

ICER: Incremental Cost-Effectiveness Ratio

Threshold = $5,180 USD. Market exchanged rate 2019: $1 USD = $3,281 COP

**Appendix F. Figure 1.** Ocrelizumab vs rituximab cost-effectiveness plane

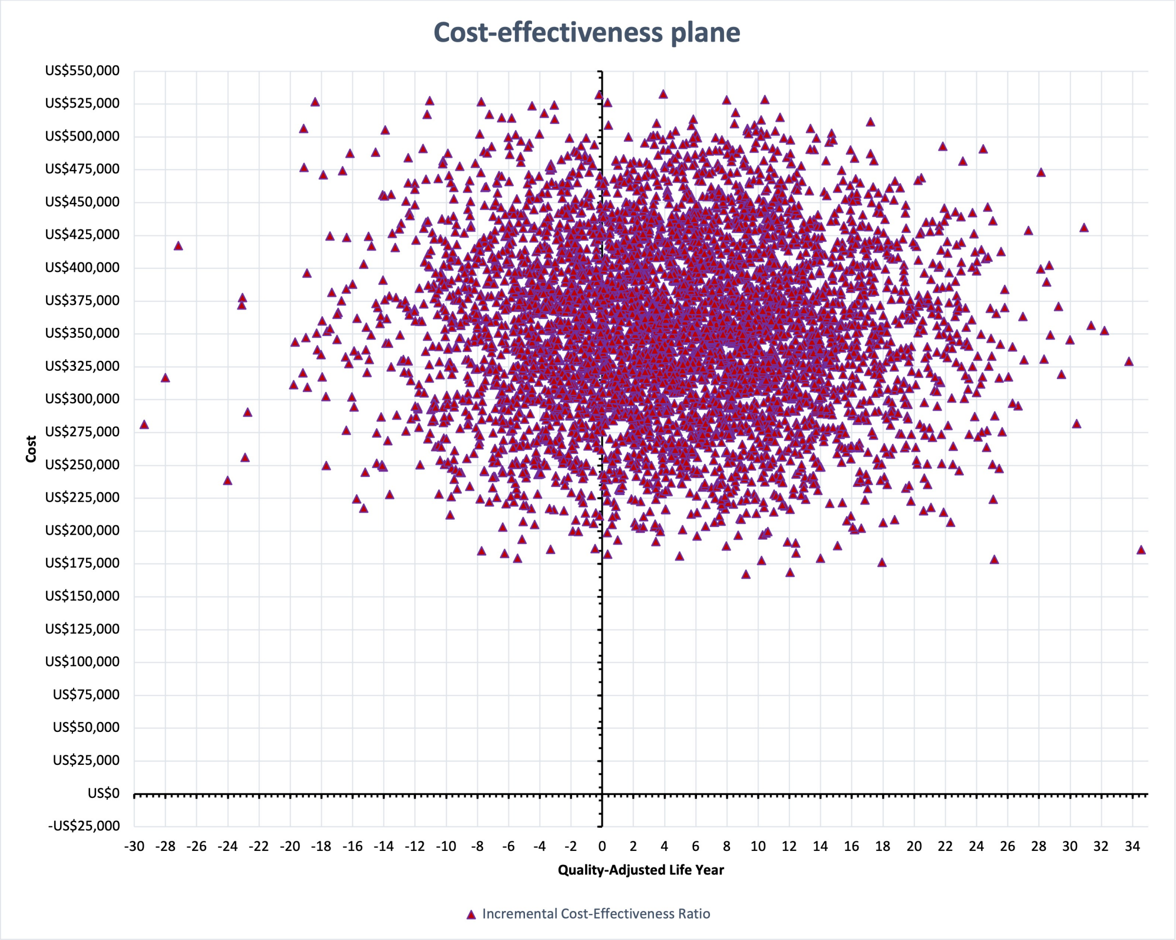

**Appendix G. Figure 2.** Ocrelizumab acceptability curve

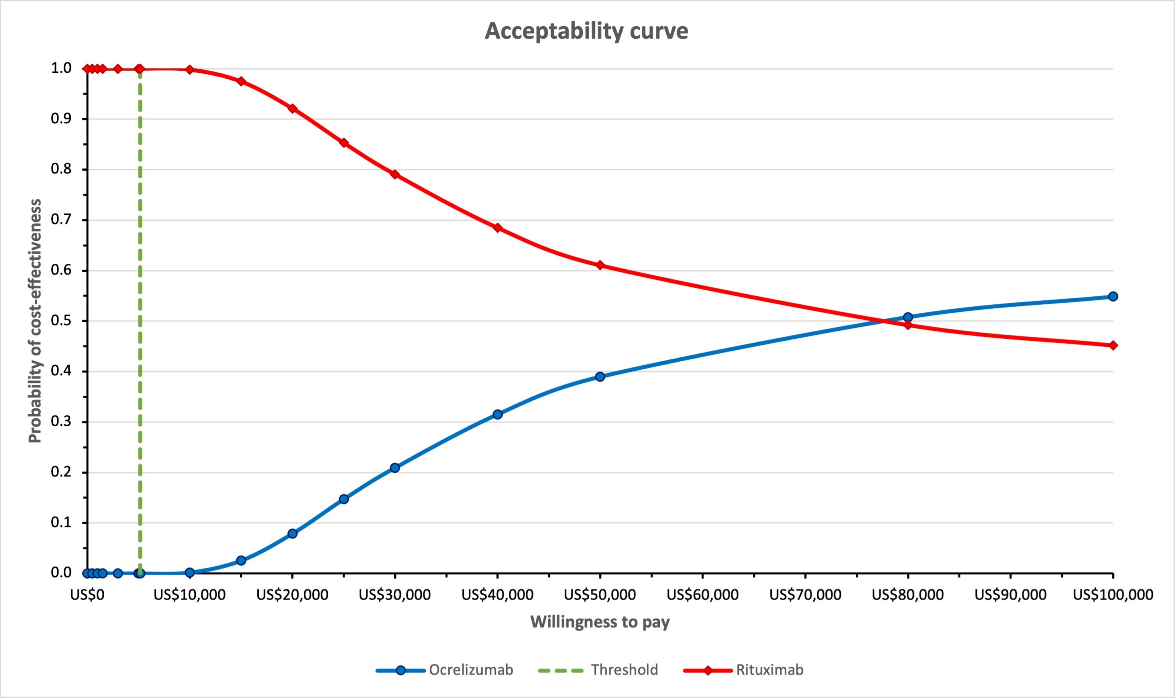
