## Supplementary material for "Cost-utility analysis comparing ocrelizumab vs rituximab in the treatment of relapsing-remitting multiple sclerosis: The Colombian perspective": CHEERS checklist

### CHEERS 2022 Checklist

| | Item | Guidance for Reporting | Reported in section |
|---|---|---|---|
| **TITLE** | | | |
| Title | 1 | Identify the study as an economic evaluation and specify the interventions being compared. | yes |
| **ABSTRACT** | | | |
| Abstract | 2 | Provide a structured summary that highlights context, key methods, results and alternative analyses. | yes |
| **INTRODUCTION** | | | |
| Background and objectives | 3 | Give the context for the study, the study question and its practical relevance for decision making in policy or practice. | lines 5-41 |
| **METHODS** | | | |
| Health economic analysis plan | 4 | Indicate whether a health economic analysis plan was developed and where available. | lines 45-54 |
| Study population | 5 | Describe characteristics of the study population (such as age range, demographics, socioeconomic, or clinical characteristics). | lines 58-60 |
| Setting and location | 6 | Provide relevant contextual information that may influence findings. | line 51 |
| Comparators | 7 | Describe the interventions or strategies being compared and why chosen. | lines 47-50 |
| Perspective | 8 | State the perspective(s) adopted by the study and why chosen. | line 51 |
| Time horizon | 9 | State the time horizon for the study and why appropriate. | line 59 |
| Discount rate | 10 | Report the discount rate(s) and reason chosen. | lines 53-55 |
| Selection of outcomes | 11 | Describe what outcomes were used as the measure(s) of benefit(s) and harm(s). | line 52 |
| Measurement of outcomes | 12 | Describe how outcomes used to capture benefit(s) and harm(s) were measured. | line 84 |
| Valuation of outcomes | 13 | Describe the population and methods used to measure and value outcomes. | line 84 |
| Measurement and valuation of resources and costs | 14 | Describe how costs were valued. | lines 90-100 |
| Currency, price date, and conversion | 15 | Report the dates of the estimated resource quantities and unit costs, plus the currency and year of conversion. | line 99 |
| Rationale and description of model | 16 | If modelling is used, describe in detail and why used. Report if the model is publicly available and where it can be accessed. | line 45 |
| Analytics and assumptions | 17 | Describe any methods for analysing or statistically transforming data, any extrapolation methods, and approaches for validating any model used. | lines 120-127 |
| Characterizing heterogeneity | 18 | Describe any methods used for estimating how the results of the study vary for sub-groups. | lines 120-127 |
| Characterizing distributional effects | 19 | Describe how impacts are distributed across different individuals or adjustments made to reflect priority populations. | not applicable |
| Characterizing uncertainty | 20 | Describe methods to characterize any sources of uncertainty in the analysis. | lines 120-127 |
| Approach to engagement with patients and others affected by the study | 21 | Describe any approaches to engage patients or service recipients, the general public, communities, or stakeholders (e.g., clinicians or payers) in the design of the study. | not applicable |
| **RESULTS** | | | |
| Study parameters | 22 | Report all analytic inputs (e.g., values, ranges, references) including uncertainty or distributional assumptions. | line 117 |
| Summary of main results | 23 | Report the mean values for the main categories of costs and outcomes of interest and summarise them in the most appropriate overall measure. | lines 136-148 |
| Effect of uncertainty | 24 | Describe how uncertainty about analytic judgments, inputs, or projections affect findings. Report the effect of choice of discount rate and time horizon, if applicable. | lines 154-175 |
| Effect of engagement with patients and others affected by the study | 25 | Report on any difference patient/service recipient, general public, community, or stakeholder involvement made to the approach or findings of the study | not applicable |
| **DISCUSSION** | | | |
| Study findings, limitations, generalizability, and current knowledge | 26 | Report key findings, limitations, ethical or equity considerations not captured, and how these could impact patients, policy, or practice. | lines 178-255 |
| **OTHER RELEVANT INFORMATION** | | | |
| Source of funding | 27 | Describe how the study was funded and any role of the funder in the identification, design, conduct, and reporting of the analysis | yes |
| Conflicts of interest | 28 | Report authors conflicts of interest according to journal or International Committee of Medical Journal Editors requirements. | yes |

Husereau D, Drummond M, Augustovski F, de Bekker-Grob E, Briggs AH, Carswell C, Caulley L, Chaiyakunapruk N, Greenberg D, Loder E, Mauskopf J, Mullins CD, Petrou S, Pwu RF, Staniszewska S; CHEERS 2022 ISPOR Good Research Practices Task Force. Consolidated Health Economic Evaluation Reporting Standards 2022 (CHEERS 2022) Statement: Updated Reporting Guidance for Health Economic Evaluations. BMJ. 2022;376:e067975.
The checklist is Open Access distributed in accordance with the terms of the Creative Commons Attribution (CC BY 4.0) license, which permits others to distribute, remix, adapt and build upon this work, for commercial use, provided the original work is properly cited. See: http://creativecommons.org/licenses/by/4.0/.
